# Utility of a Score-Based Approach to Liver Assessment in Heart Transplant Candidates

**DOI:** 10.1101/2023.05.19.23290258

**Authors:** Joshua A. Rushakoff, Louie Cao, Joe Ebinger, Alexander Kuo, Patrick Botting, Dominic Emerson, Guillame Countance, Pascal Lebray, Rose Tompkins, Jon A. Kobashigawa, Jignesh K. Patel, Maha Guindi, Evan P. Kransdorf

## Abstract

**Background:** While abnormalities of liver function and imaging are common in patients with end-stage heart failure, advanced fibrosis is uncommon. Liver biopsy (LB) is used to identify advanced fibrosis in heart transplant (HT) candidates but can delay or limit access to definitive therapies and cause complications. We sought to develop and determine the utility of a clinical risk score for advanced fibrosis in HT candidates.

**Methods:** We conducted a retrospective, single-center review of patients evaluated for HT between 2012 and 2019 (n = 1,651) and identified those who underwent LB (n = 137) as well as a matched control cohort (n = 160). All biopsies were reviewed by a liver pathologist. Kaplan-Meir curves were used to assess survival. Univariate logistic modeling was used to identify factors predictive of advanced liver fibrosis. Simulation using synthetic data bootstraps was performed to determine the utility of using a score-based approach to trigger LB.

**Results:** We identified 32 (23%) patients with stage 0, 79 (58%) with stage 1-2, and 26 (19%) with stage 3-4/advanced fibrosis. We found no difference in survival at 3 years post-HT based on pre-HT fibrosis stage. The factor most associated with pursuit of LB was abnormal liver parenchyma on ultrasound. We found that a score combining severe tricuspid regurgitation, alcohol use, and low-density lipoprotein improved specificity and reduced the number of LBs required.

**Conclusions:** A score composed of non-invasive factors may help reduce the number of patients who require LB for diagnosis of advanced fibrosis. Additional multicenter studies are needed to validate this score.

## 1. Introduction

Liver dysfunction is common in the setting of end stage heart failure ^1–3^. Heart transplant (HT) candidates with abnormal liver laboratory studies or abnormal liver parenchyma by imaging warrant further evaluation as passive hepatic congestion due to chronically elevated right heart pressures can lead to irreversible sinusoidal dilation, centrilobular necrosis, and progressive fibrosis ^3, 4^, which is a risk factor for poor post-HT outcome. Therefore, assessment of liver fibrosis in HT candidates is of paramount importance as the degree of fibrosis may determine lone-HT candidacy or dictate the need for combined heart liver transplant ^5^.

Multiple clinical risk scores including combining liver biopsy (LB) stage with Model for End Stage Liver Disease Excluding INR (MELD-XI) ^6^ and combining the Model for End Stage Liver Disease (MELD) with ascites ^7^ have been proposed to predict liver fibrosis stage in the setting of congestive hepatopathy (CH) with mixed results. Others have evaluated the utility of biomarkers including total bilirubin and albumin ^4, 8, 9^ for prediction of liver fibrosis with variable benefit in CH. Further, in Fontan patients imaging modalities including ultrasound, computed tomography, magnetic resonance ^10, 11^, transient elastography ^12^ and laboratory studies ^13, 14^ have proved inconsistent in predicting liver fibrosis. In non-CH cirrhosis, other scores including the Lok Index, King Score, aspartate aminotransferase (AST) to platelet ratio index (APRI), FIB4, and AST to alanine aminotransferase (ALT) ratio (AAR), have been shown to be predictive of cirrhosis, although these have not been thoroughly evaluated in patients with CH ^15, 16^. The poor performance of previously proposed biochemical markers and clinical risk scores has led to a reliance on LB for HT candidate evaluation ^1, 5^. The LB has important limitations. First, the need to obtain a LB can delay or limit access (when it is not available) to definitive therapies such as implantation of mechanical circulatory support or HT. Next, LB-explant comparison series in patients with CH have shown heterogeneity of fibrosis, making LB interpretation complex ^9, 17^. Furthermore, evidence has emerged that fibrosis stage on LB does not predict post-HT survival ^9, 17, 18^. LB is also not without risk of complications ^19^.

Clinical risk scores for liver fibrosis have not been evaluated in patients with CH and many of the previous studies on CH have been focused on Fontan patients. As such, we sought to identify non-invasive factors that predict advanced fibrosis and assess the ability of clinical risk scores to predict advanced fibrosis in a population of non-congenital HT candidates. We also sought to determine if a score-based approach could improve identification of candidates at risk of having advanced liver fibrosis requiring LB.

## 2. Methods and Materials

This study was performed under an approved protocol of the Cedars-Sinai Institutional Review Board (STUDY00002007).

### Patient Selection

All adult patients who were evaluated for HT at Cedars-Sinai Medical Center between 1/1/2012 and 12/31/2019 (n=1,651) were reviewed (**Figure 1**). Those patients with a pre-transplant LB (n = 167) were identified. A control cohort was developed by selecting at random an equal number of candidates that underwent HT in the same year but that did not undergo pre-transplant LB (n=167). Candidates with congenital heart disease and those with inadequate (less than 1.5 cm in length) /unavailable biopsies or absent laboratory studies/echocardiograms were excluded yielding a final cohort of 137 HT candidates with LB and 160 HT candidates without LB.

**Figure 1.**
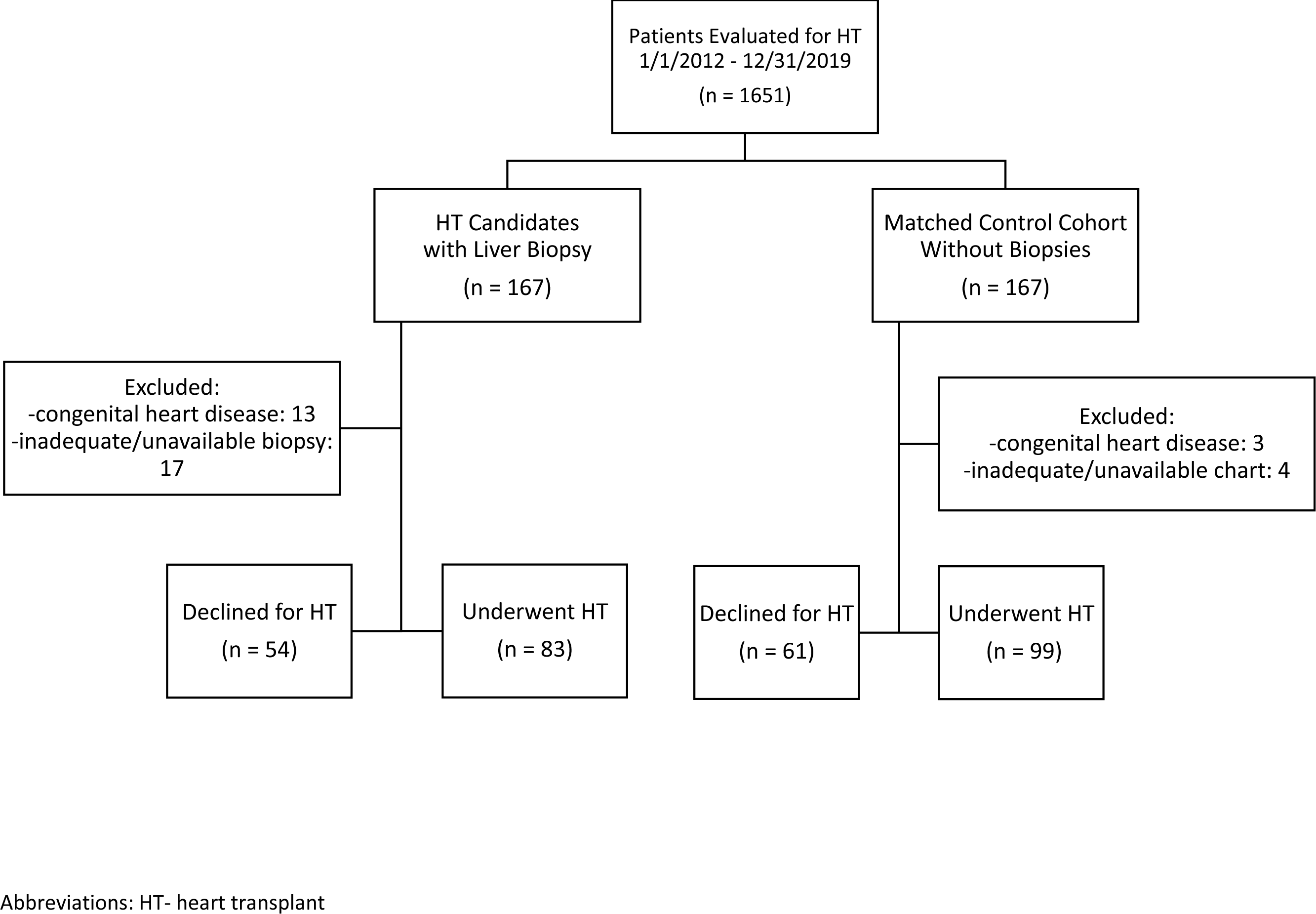
Cohort Construction.

All candidates were evaluated by a heart failure/transplant cardiologist and their candidacy evaluated in a multidisciplinary meeting. If grade 3 or 4 fibrosis was identified by LB candidates were also evaluated by a transplant hepatologist. Selection criteria for heart transplant were consistent with ISHLT guidelines ^5^. Selection criteria for combined heart liver transplant (CHLT) were as for heart alone but with predominance of grade 4 fibrosis on LB and age:<S 65 years. In the candidates with LB, 8 received CHLT (all with advanced fibrosis). In the candidates without LB, 1 received CHLT (history of hepatitis C cirrhosis).

### Biopsies and Histologic Assessment

Liver biopsies were retrieved from our pathology warehouse. For patients with multiple biopsies, the biopsy closest to HT was reviewed. All LB were obtained by interventional radiologists. 126 biopsies were obtained via a transjugular approach with an 18-gauge needle (median number of passes = 4) and 11 biopsies via a percutaneous ultrasound guided approach.

Biopsies were scored from stage 0 to stage 4 based on a fibrosis staging system for CH (stage 0 - no fibrosis; stage 1 - zone 3 fibrosis; stage 2A - zone 3 and mild portal fibrosis with accentuation at zone 3; stage 2B - at least moderate portal fibrosis and zone 3 fibrosis with accentuation at portal zone; stage 3 - bridging fibrosis; and stage 4 - cirrhosis) ^18, 20^. Nodular regenerative hyperplasia (NRH) was assessed in non-fibrotic areas on H&E and reticulin stains and considered present if involving more than 50 percent of the biopsy. Stage 3 and stage 4 fibrosis were considered advanced fibrosis. All biopsies were reviewed by a single expert liver pathologist (M.G.).

### Clinical Variables

Candidate characteristics at the time of HT evaluation including review of transthoracic echocardiograms (TTE), right heart catheterizations, laboratory studies, and abdominal ultrasounds. Components of previously developed risk scores King score^21^, APRI ^22^, Lok Index^23^, FIB-4 Score^24^, AAR^25^, Columbia Liver Risk Score^6^, MELD-XI ^26^, and Varices-Ascites-Splenomegaly-Thrombocytopenia (VAST) ^27^ were collected **(Supplementary Table 1).** The minimum value for each laboratory variable during the pre-transplant period was utilized for the calculation of risk scores. The median time between laboratory variables and transplant or LB is provided in **Supplementary Table 2.**

### Statistical Analysis

Liver fibrosis stage was separated into 3 groups based on the predominant fibrosis stage on the biopsy: group 1: no fibrosis/stage 0, group 2: mild fibrosis/stages 1-2, group 3: advanced fibrosis/stages 3-4. Variables/factors assessed included: 1) echocardiographic factors as graded by initial reader: ejection fraction (EF), right ventricular (RV) dysfunction grade, mitral regurgitation (MR) grade, tricuspid regurgitation (TR) grade, left ventricular internal diameter end diastole (LVIDd), 2) cardiac hemodynamic factors: right atrial pressure, pulmonary pressures, pulmonary capillary wedge pressure, ratio of right atrial pressure to pulmonary capillary wedge pressure, 3) hepatic hemodynamic factors: wedged hepatic venous pressure, free hepatic venous pressure, hepatic venous pressure gradient, 4) blood chemistry factors: sodium, serum creatinine, total bilirubin, serum albumin, ALT, AST, platelet count, LDL (low-density lipoprotein), triglycerides, hepatitis B core antibody, and hepatitis C antibody, 5) abdominal ultrasound factors: liver size, liver parenchymal appearance, presences of ascites (at a level greater than “trace”), and 6) demographic factors: age, sex, and history of heavy alcohol use (men: ≥ 4 drinks on any day or ≥ 14 drinks per week; women: ≥ 3 drinks on any day or ≥ 7 drinks per week) ^28^.

Baseline characteristics were compared using analysis of variance (ANOVA) for continuous normally-distributed variables, Kruskall-Wallis test for continuous non-normally-distributed variables, and chi-squared test for categorical variables implemented in the R package “compareGroups” ^29^. Survival modeling was performed with death at 3 years as the response variable using Kaplan-Meier method and Cox models implemented in the R package “survival” ^30^. Performance of previously identified scores was assessed through calculation of area under the receiver operating characteristic curve (AUROC) with advanced fibrosis as the response variable.

In the set of candidates that underwent LB, we performed univariable logistic modeling to determine factors associated with performance of a LB and factors associated with advanced fibrosis. Given that there were only 26 candidates with advanced fibrosis, we were limited in the number of variables that could be assessed in multivariable modeling. Significant (p ý 0.05) variables in the univariate analysis were included in a multivariable logistic model. Prior to multivariable analysis missing values for LDL (n=20), MR (n=6), TR (n=5), and US (n=2) were imputed using the R package “mice” ^31^.

We performed simulations using synthetic data bootstraps to determine the utility of using a score-based approach to trigger LB. We generated a clinical risk score for advanced fibrosis by assigning points using the beta10/integer method to variables with p ý 0.05 in the multivariable logistic model **(Supplementary Table 3**). Simulation 1: For the set of candidates that did not undergo LB, we used variables with p ý 0.05 in the multivariable logistic model to predict the risk of advanced fibrosis, using a predicted probability of ≥ 0.5 as indicative of advanced fibrosis. This resulted in 6 of 160 candidates having advanced fibrosis. These candidates were combined with candidates that did undergo LB for simulations. Missing values were imputed as above. In each of 1000 bootstraps, we generated a synthetic cohort using the R package “synthpop” ^32^. Clinical risk score was calculated for each synthetic candidate. The optimal cutpoint for the score was determined using the F1-score implemented in the R package “cutpointr” ^33^. LB was considered to be indicated: 1) when the clinical risk score was 2 optimal cutpoint or 2) if the US showed a nodular parenchyma. The total number and percent of LB required for the synthetic cohort was tallied. Area under the receiver operating characteristic curve (AUROC), sensitivity and specificity of each biopsy approach for the diagnosis of advanced liver disease was calculated. Simulation 2: The median optimal cutpoint for the clinical risk score derived from simulation 1 was -4. We repeated the simulation above with the same conditions except that we used a cutpoint of -4 for all bootstraps and performance of each biopsy approach for the diagnosis of advanced liver disease was calculated.

All analyses were completed in R version 4.0.5 (The R Foundation for Statistical Computing).

## 3. Results

### Baseline Characteristics

The cohort of patients was grouped based on the predominant fibrosis stage on LB (**Table 1**). We identified 32 (23%) patients with no fibrosis, 79 (58%) with mild fibrosis (stages 1-2), and 26 (19%) with advanced fibrosis (stages 3-4). There were no significant differences in baseline demographic factors including age, gender, comorbidities, and presence of durable left ventricular assist device. A higher percentage of patients with advanced fibrosis (31%) had a history of heavy alcohol use compared to patients with no fibrosis (9%) or mild fibrosis (8%). As expected, fewer patients with advanced fibrosis (19%) underwent lone-HT compared to those with mild fibrosis (67%), no fibrosis (53%), or the no biopsy control group (61%).

**Table 1.** Baseline Characteristics.

|  | Predominant Fibrosis Stage on Biopsy |  |  |  |  |
| --- | --- | --- | --- | --- | --- |
|  | No Biopsy<br>(n = 160) | No Fibrosis<br>(n = 32) | Mild Fibrosis<br>(stages 1-2)<br>(n = 79) | Advanced Fibrosis<br>(stages 3-4)<br>(n = 26) | p-value |
| **Demographics** |  |  |  |  |  |
| Age (years) | 57.1 (11.0) | 57.8 (11.3) | 56.6 (10.7) | 59.0 (9.65) | 0.774 |
| Male | 120 (75.0%) | 25 (78.1%) | 69 (87.3%) | 21 (80.8%) | 0.176 |
| NICM | 80 (50.0%) | 21 (65.6%) | 50 (63.3%) | 20 (76.9%) | 0.070 |
| Diabetes Mellitus | 62 (38.8%) | 9 (28.1%) | 25 (32.1%) | 6 (23.1%) | 0.315 |
| Hypertension | 69 (43.1%) | 10 (31.2%) | 27 (34.6%) | 9 (34.6%) | 0.428 |
| BMI (kg/m<sup>2</sup>) | 25.9 (5.7%) | 26.7 (5.18) | 25.9 (4.89) | 25.8 (4.98) | 0.878 |
| Left Ventricular Assist Device | 28 (17.8%) | 12 (37.5%) | 14 (17.9%) | 4 (15.4%) | 0.066 |
| History of heavy alcohol use | 11 (6.9%) | 3 (9.4%) | 6 (7.6%) | 8 (30.8%) | 0.007 |
| Hepatitis C Positive | 4 (2.8%) | 0 (0.00%) | 3 (4.1%) | 2 (8.3%) | 0.320 |
| Hepatitis B Positive | 26 (10.3%) | 18 (13.2%) | 1 (3.5%) | 5 (7.4%) | 0.378 |
| **Abdominal Ultrasound Findings** |  |  |  |  |  |
| Ascites | 33 (25.0%) | 16 (51.6%) | 43 (54.4%) | 18 (72.0%) | <0.001 |
| Splenomegaly | 19 (14.4%) | 9 (29.0%) | 22 (27.8%) | 9 (36.0%) | 0.021 |
| Liver Parenchyma |  |  |  |  | <0.001 |
| Heterogenous | 33 (25.0%) | 8 (25.8%) | 26 (32.9%) | 8 (32.0%) |  |
| Nodular | 6 (4.55%) | 12 (38.7%) | 35 (44.3%) | 13 (52.0%) |  |
| Liver Size |  |  |  |  |  |
| Large | 35 (26.5%) | 15 (48.4%) | 22 (27.8%) | 11 (44.0%) | 0.009 |
| **Portal Pressure Measurements** |  |  |  |  |  |
| Free Hepatic Venous Pressure (mmHg) |  | 9.85 (5.85) | 13.5 (7.98) | 15.4 (7.04) | 0.024 |
| Hepatic Wedge Pressure (mmHg) |  | 13.8 (7.68) | 17.0 (8.17) | 19.4 (7.90) | 0.051 |
| **Transthoracic Echocardiogram** |  |  |  |  |  |
| Ejection Fraction (%) | 22.7 (11.6) | 22.5 (10.2) | 27.8 (16.9) | 26.0 (13.3) | 0.041 |
| Right Ventricular Function |  |  |  |  |  |
| Severely Depressed | 21 (14.6%) | 11 (36.7%) | 17 (23.0%) | 6 (27.3%) |  |
| Mitral Regurgitation |  |  |  |  | 0.014 |
| Mild | 41 (28.7%) | 7 (23.3%) | 20 (26.3%) | 3 (12.0%) |  |
| Moderate | 29 (20.3%) | 13 (43.3%) | 29 (38.2%) | 9 (36.0%) |  |
| Severe | 36 (25.2%) | 8 (26.7%) | 14 (18.4%) | 10 (40.0%) |  |
| Tricuspid Regurgitation |  |  |  |  | <0.001 |
| Mild | 38 (26.0%) | 7 (23.3%) | 20 (26.0%) | 3 (12.0%) |  |
| Moderate | 39 (26.7%) | 11 (36.7%) | 28 (36.4%) | 3 (12.0%) |  |
| Severe | 31 (21.2%) | 7 (23.3%) | 21 (27.3%) | 16 (64.0%) |  |
| LVIDD (cm) | 3.12 (0.72) | 3.18 (0.67) | 2.99 (0.77) | 3.29 (0.68) | 0.285 |
| IVC Collapsibility |  |  |  |  | <0.001 |
| None (RA pressure ~15) | 39 (30.2%) | 13 (48.1%) | 43 (56.6%) | 15 (65.2%) |  |
| <50 % (RA Pressure ~8) | 39 (30.2%) | 8 (29.6%) | 24 (31.6%) | 6 (26.1%) |  |
| Right Heart Catheterization Pressures |  |  |  |  |  |
| RAP (mmHg) | 8.85 (5.34) | 11.3 (5.83) | 14.9 (6.62) | 15.2 (7.56) | <0.001 |
| PAs (mmHg) | 43.8 (14.1) | 44.7 (11.6) | 48.2 (14.1) | 41.1 (11.6) | 0.105 |
| PCWP (mmHg) | 20.0 (8.84) | 20.9 (9.20) | 23.7 (8.82) | 21.0 (8.40) | 0.065 |
| RAP to PCWP Ratio | 0.48 (0.26) | 0.55 (0.22) | 0.66 (0.30) | 0.72 (0.31) | <0.001 |
| Laboratory Values |  |  |  |  |  |
| Albumin (g/dL) | 2.99 (0.57) | 2.85 (0.62) | 2.90 (0.51) | 3.12 (0.57) | 0.219 |
| Total Bilirubin (mg/dL) | 1.16 (0.57) | 1.62 (1.59) | 1.11 (0.41) | 1.06 (0.24) | 0.003 |
| Serum Creatinine (mg/dL) | 0.90 (0.44) | 0.90 (0.46) | 1.00 (0.50) | 1.14 (1.05) | 0.140 |
| Platelet Count (x10<sup>9</sup>/L) | 91.0 (53.6) | 78.4 (52.3) | 78.8 (32.1) | 89.0 (48.1) | 0.227 |
| Sodium (mmol/L) | 129 (4.41) | 128 (3.78) | 128 (4.62) | 128 (6.07) | 0.078 |
| Alanine Aminotransferase (units/L) | 16.8 (15.4) | 14.9 (11.4) | 12.6 (6.43) | 13.1 (7.60) | 0.086 |
| Aspartate Aminotransferase (units/L) | 23.2 (35.5) | 23.5 (22.9) | 16.9 (6.57) | 16.2 (7.87) | 0.282 |
| LDL (mg/dL) | 62.6 (25.9) | 69.1 (28.5) | 61.1 (22.0) | 48.7 (17.0) | 0.029 |
| Triglyceride (mmol/L) | 92.0 (51.5) | 109 (92.3) | 68.8 (27.3) | 81.5 (40.8) | 0.002 |
| Patients who underwent transplant |  |  |  |  |  |
| UNOS Status at Listing |  |  |  |  | 0.306 |
| Status 1-2 | 17 (18.7%) | 3 (21.4%) | 7 (15.6%) | 0 |  |
| Status 3 | 51 (56.0%) | 9 (64.3%) | 33 (73.3%) | 5 (100%) |  |
| Status 4-6 | 23 (25.3%) | 2 (14.3%) | 5 (11.1%) | 0 |  |
| Inotropes at Listing | 34 (37.4%) | 5 (35.7%) | 28 (62.2%) | 3 (60.0%) | 0.030 |

Abbreviations: NICM: non-ischemic cardiomyopathy; BMI- body mass index; LVIDD – left ventricular internal diameter end diastole; IVC – inferior vena cava; RAP- right atrium pressure; PAs- pulmonary artery systolic; PCWP- pulmonary capillary wedge pressure

There were significant differences between the groups in the presence of ascites (p <0.001) and splenomegaly (p = 0.02), with these most common in the advanced fibrosis group. Liver parenchyma on ultrasound was also significantly different between the groups (p <0.001) with nodularity most common in the advanced fibrosis group (52%). Finally, hepatic venous pressure measurements at the time of LB showed stepwise increases with increasing fibrosis stage.

Interestingly, valvular disease was more common in the advanced fibrosis group (**Table 1**). 40% of the advanced fibrosis cohort had severe MR, compared to 25% of the no-biopsy group, 27% of the biopsy no fibrosis group, and 18% of the mild fibrosis group (p = 0.014). Similarly, 64% of the advanced fibrosis group had severe TR compared to 21%, 23%, and 27% of the no-biopsy, biopsy-no fibrosis, and mild fibrosis groups, respectively. These hemodynamic impacts were also observed on right heart catheterization with significant differences in right atrial pressure (p<0.001) and right atrial pressure to pulmonary capillary wedge pressure ratio (p<0.001). While laboratory markers were largely similar across the groups, we did observe that the advanced fibrosis group had the lowest total bilirubin (p=0.003) and LDL (p=0.029). For those who underwent transplant, UNOS listing status was similar across groups while inotrope use at time of transplant was more common in the mild fibrosis and advanced fibrosis groups (p=0.030).

### Factors Influencing Obtaining Biopsy and Utility of Ultrasound

Using univariate logistic modeling we identified factors associated with obtaining a LB during the pre-HT period **(Supplementary Table 5)**. Most notable were nodular liver parenchyma on abdominal ultrasound (OR 7.48, 95% CI 2.33-24.01, p= 0.001), heterogenous liver parenchyma on abdominal ultrasound (OR 3.64, 95% CI 1.06-12.54, p =0.041), and presence of ascites on abdominal ultrasound (OR 4.19, 95% CI 1.69-10.42, p=0.002). Additionally, severe valvular disease was associated with obtaining a LB (severe MR [OR 3.91, 95% CI 1.03-14.88, p = 0.046], severe TR [OR 5.88, 95% 1.63-21.19, p = 0.007]). Given the strong odds of obtaining a LB in a patient with a nodular liver parenchyma, we then analyzed the performance of ultrasound parenchyma in predicting advanced fibrosis and found poor predictive ability (AUROC 0.566) (**Table 2**).

**Table 2.** Performance of abdominal ultrasound and extant non-invasive scoring systems in predicting advanced fibrosis stage.

|  | **AUC (95% CI)** | **Sensitivity** | **Specificity** | **Accuracy** |
| --- | --- | --- | --- | --- |
| Abdominal Ultrasound: Nodular Liver | 0.566 (0.526 – 0.784) | 0.560 | 0.573 | 0.570 |
| Lok Index | 0.528 (0.369 – 0.688) | 0.563 | 0.582 | 0.578 |
| King Score | 0.542 (0.403 – 0.681) | 0.346 | 0.938 | 0.479 |
| APRI | 0.638 (0.493 – 0.782) | 0.346 | 0.938 | 0.479 |
| FIB4 | 0.578 (0.422 – 0.735) | 0.291 | 0.875 | 0.423 |
| AAR | 0.562 (0.387 – 0.737) | 0.500 | 0.764 | 0.704 |
| MELDXI | 0.529 (0.369 - 0.690) | 0.673 | 0.500 | 0.634 |
| VAST | 0.677 (0.552 - 0.794) | 0.944 | 0.333 | 0.493 |

Abbreviations: APRI- AST to platelet ratio index; AAR- AST to ALT ratio; MELDXI- model for end stage liver disease excluding INR; VAST – varices, ascites, splenomegaly, thrombocytopenia

### Biopsy Pathology and Post-HT Survival

We next calculated post-HT survival based on the predominant fibrosis stage on the pre-HT LB. Candidates that underwent combined heart-liver transplant (n=9) were excluded from this analysis. Notably, there was no difference in 3-year post-HT survival between the groups (**Figure 2**). NRH, another histologic marker of liver damage, can be identified in patients with CH without cirrhosis. Excluding patients with advanced fibrosis, we found NRH to be common (28% biopsy-no fibrosis, 37% biopsy-mild fibrosis) in this sub-population. However, we saw no difference in 3-year survival based on the presence of NRH **(Supplementary Figure 1).**

**Figure 2.**
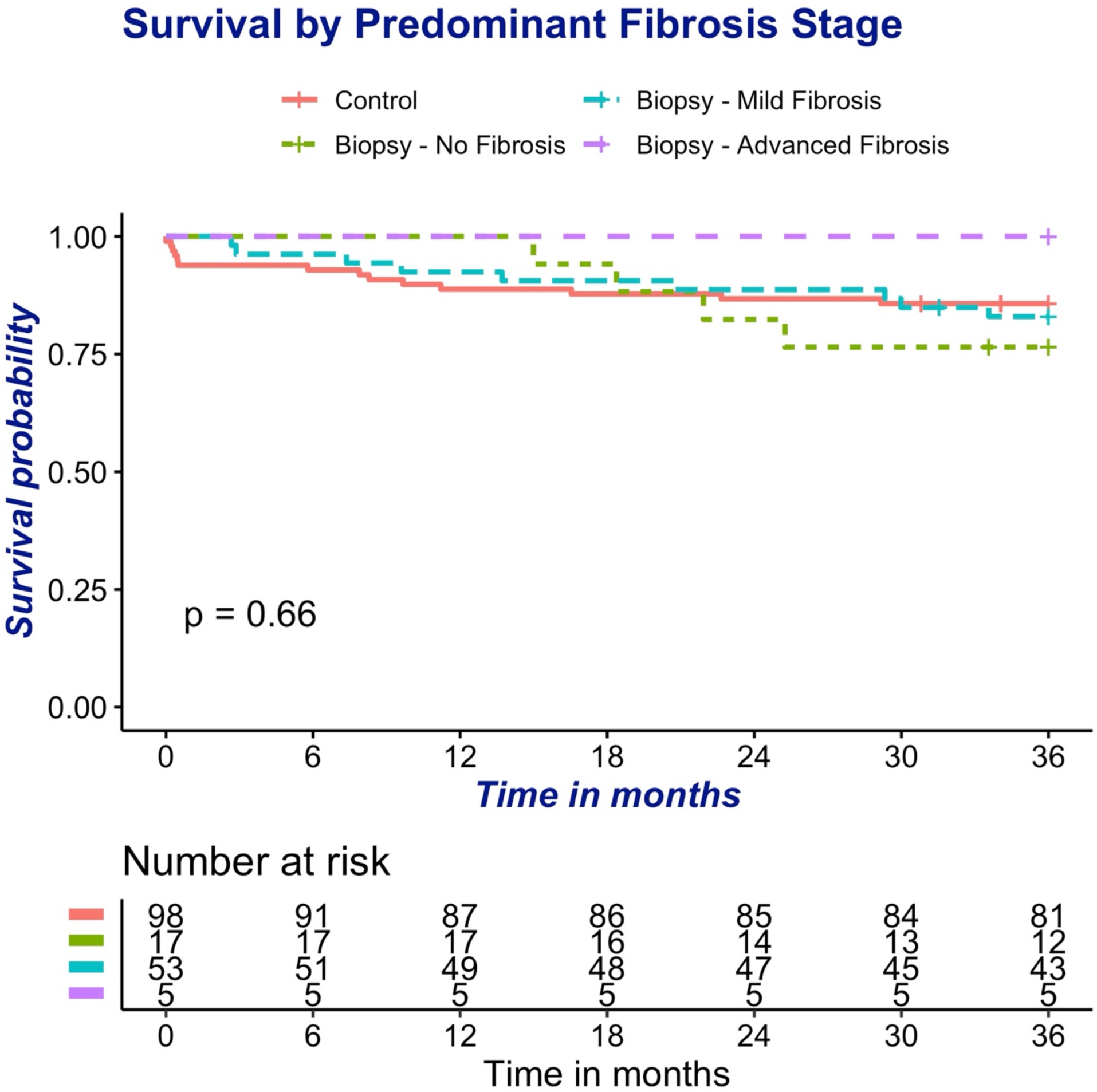
**Kaplan Meir Survival Curve by Fibrosis Stage**.

### Prediction Score Analysis

Next, we assessed the performance of previously proposed non-invasive scoring systems for liver fibrosis. AUROC for prediction of advanced fibrosis stage was calculated for the Lok index (0.472), King score (0.443), APRI (0.606), FIB4 (0.465), AAR (0.519), MELD-XI (0.529), and VAST (0.677) scores (**Table 2**). Additionally, we assessed the previously proposed Columbia score which was developed in a similar population of HT candidates ^6^. Comparing patients below and at or above the predefined cutoff of 45 by the Columbia Liver Risk Score, we found no difference in survival between the groups (**Supplementary Figure 2)**.

### Univariate Analysis and Creating of Predictive Score for Advanced Fibrosis

For patients who underwent LB, demographic, hemodynamic, echocardiographic, and blood chemistry factors were evaluated with univariate logistic modeling to assess the ability to predict advanced fibrosis **(Supplementary Table 6).** A history of heavy alcohol use (OR 5.56, 95% CI 2.15-14.35, p < 0.001) and severe valvular disease was associated with increased odds of advanced fibrosis (severe MR [OR 2.55, 95% CI 1.01-6.44, p = 0.048], severe TR [OR 5.02, 95% CI 1.99-12.63, p < 0.001]). Low-density lipoprotein (LDL) was the only laboratory marker predictive of advanced fibrosis (OR 0.97 per mg/dL, 95% CI 0.94-0.99, p = 0.009). Of non-invasive scores, the VAST (OR 1.73 per point, 95% CI 0.95-3.12, p = 0.070) exhibited a trend toward statistical significance. Additional factors that exhibited a trend towards an increased odds of advanced fibrosis included ascites on abdominal ultrasound (OR 2.22, 95% CI 0.86-5.75, p =0.099) and serum albumin (OR 2.22 per mg/dL, 95% CI 0.97-5.09, p=0.060). Notably, nodular liver parenchyma on abdominal ultrasound was not associated with increased odds of advanced fibrosis (OR 2.01, 95% CI 0.60-6.74, p = 0.261).

Next, factors with p ý 0.05 in univariate modeling were added to a multivariable logistic model (**Table 3**). Severe TR (OR 3.23, 95% CI 1.93-5.39, p = 0.023), a history of heavy alcohol use (OR 4.62, 95% CI 2.52-8.45, p = 0.011), and LDL (OR 0.96 per mg/dL, 95% CI 0.95-0.98) remained predictive of advanced fibrosis in the multivariable model.

**Table 3.** Multivariable Analysis of Factors for Advanced Fibrosis.

|  | Odds Ratio | 95% Confidence Intervals | p-value |
| --- | --- | --- | --- |
| Severe TR | 3.23 | 1.93-5.39 | 0.023 |
| Severe MR | 1.25 | 0.73-2.14 | 0.682 |
| History of heavy alcohol use | 4.62 | 2.52-8.45 | 0.011 |
| LDL | 0.96 | 0.95-0.98 | 0.011 |

Abbreviations: TR- tricuspid regurgitation; MR- mitral regurgitation; LDL- low-density lipoprotein

**Table 4.** Evaluation of Predictive Score for Advanced Fibrosis in Synthetic Dataset.

|  | AUROC | Sensitivity | Specificity | Percent Liver Biopsies Required |
| --- | --- | --- | --- | --- |
| *Simulation 1: Using optimal cutpoint identified for each bootstrap* |  |  |  |  |
| Score: Severe TR + Heavy Alcohol Use + LDL | 0.71<br>(0.63-0.80) | 48%<br>(33%-76%) | 93%<br>(78-99%) | 10%<br>(4%-25%) |
| US only | 0.50<br>(0.43-0.58) | 41%<br>(22%-100%) | 73%<br>(0%-81%) | 23%<br>(18%-28%) |
| *Simulation 2: Using cutpoint of -4 for all bootstraps* |  |  |  |  |
| Score: Severe TR + Heavy Alcohol Use + LDL | 0.69<br>(0.59-0.78) | 44%<br>(26%-62%) | 93%<br>(90-96%) | 11%<br>(7%-14%) |
| US only | 0.50<br>(0.43-0.58) | 41%<br>(22%-100%) | 73%<br>(0%-82%) | 23%<br>(18%-28%) |

Numbers represent median of all 1000 bootstraps with 95% confidence intervals in parenthesis.
Abbreviations: AUCOC - area under receiver operating characteristic curve; TR -tricuspid regurgitation; LDL- low-density lipoprotein; US - ultrasound of liver parenchyma

### Simulation of Score-Based Approach versus Ultrasound-Based Approach

We developed a clinical risk score for advanced fibrosis composed of the three significant variables in the multivariable model **(Supplementary Table 3)**. This clinical risk score was assessed in two simulations using synthetic data bootstraps. In the first simulation, using the clinical risk score (above the optimal cutpoint identified for each bootstrap) to trigger LB was associated with a significantly higher AUROC (0.71 vs. 0.50), numerically higher sensitivity (48% vs. 41%), numerically higher specificity (93% vs. 73%), and a lower percentage of LB required for the cohort (10% vs. 23%). In the second simulation, using the clinical risk score (above a single cutpoint of -4 points) we observed similar results.

## 4. Discussion

In this study, we sought to identify non-invasive factors that predict advanced fibrosis and assess the ability of clinical risk scores to predict advanced fibrosis in a population of HT candidates without congenital heart disease. We found the factor most strongly associated with performance of a LB was liver nodularity on abdominal ultrasound, whereas we found that history of heavy alcohol use and severe tricuspid regurgitation were the factors most strongly associated with advanced fibrosis in patients with CH. Existing non-invasive scores did not provide adequate performance for the prediction of advanced fibrosis. With the goal of developing a score-based approach to non-invasively identify patients most in need of a LB, we performed simulations using synthetic data bootstraps derived from our cohort and found that a score based on severe tricuspid regurgitation, heavy alcohol use, and LDL decreased the number of LBs needed to diagnose advanced fibrosis as compared to using abdominal ultrasound alone.

We found that fibrosis stage on LB did not predict survival at 3 years after HT. This was likely a consequence of our institutional protocol in which candidates for HT with predominance of grade 4 fibrosis on LB underwent heart-liver transplant; in 12 candidates with advanced fibrosis on LB that underwent HT, 7 candidates (58%) underwent heart-liver transplant. However, advanced liver fibrosis alone ^34^, or in conjunction with other clinical factors such as MELD ^6^ is a well-established risk factor for adverse outcomes after durable left ventricular assist device implantation or HT. As such, only 12 of the 26 candidates (46%) that were found to have advanced fibrosis on LB underwent HT, as compared to 64% of candidates without advanced fibrosis on LB. Thus, even though we did not identify liver fibrosis as a risk factor for post-transplant outcomes, the presence of advanced fibrosis affects the likelihood that a candidate will undergo HT and thus long-term survival.

Multiple studies have highlighted the difficulty of evaluating liver disease in end-stage heart failure. Shingara et al. recently surveyed providers at CHLT centers in North America and reported disparate evaluation and listing practices, reflecting the lack of consensus when considering these candidates ^35^. We previously reported that LB results have discordance with explanted livers in patients who underwent combined heart-liver transplants, reflecting the heterogeneity of CH and suggesting that LB in this population must be interpreted with caution ^9, 18, 36^. This finding has also been reported in Fontan patients where the liver is exposed to chronically elevated pressures ^37^. As such, there have been efforts to develop clinical risk scores to assist in evaluating the reversibility of liver damage. Most notably, Farr et al combined biopsy fibrosis stage with MELDXI and reported higher scores were predictive of 1-year post-HT mortality ^6^. We evaluated this score in our cohort and found no difference in 1 or 3-year post-HT mortality, suggesting caution in applying this score in clinical practice. Others have focused on the Fontan patient population ^38, 39^ and, notably, found the VAST score to be predictive of fibrosis ^27^. While we excluded patients with congenital heart disease to avoid potential confounding factors, we did find markers of elevated right sided pressure, like those seen in Fontan patients, were associated with advanced fibrosis. Additionally, we found lower LDL to be independently associated with advanced fibrosis, consistent with previous reporting that LDL is predictive of both survival in heart failure ^40^ and cirrhosis ^41^.

In pursuing LB, patients are exposed to potential complications. Transjugular liver biopsies have a reported major complication rate of 0.59% ^19^. While not considered specifically in previous studies, many HT candidates are on systemic anticoagulation for comorbid cardiac conditions or mechanical circulatory devices which confers an increased risk of bleeding or may preclude a LB altogether. Furthermore, in resource limited settings LB may not be readily available. These concerns highlight that the score-based approach demonstrated in this study may be of clinical utility.

Despite the challenges, accurate assessment of fibrosis stage in HT candidates is very important. Liver fibrosis stage impacts candidacy and contributes to decision making about which patients are declined for HT, are eligible for lone HT, or CHLT. In the past, those patients determined to require CHLT have experienced longer wait times and higher waitlist mortality compared to lone HT candidates ^42, 43^, though recent allocation changes may impact these trends ^44^. These adverse outcomes suggest another possible use for the score-based approach demonstrated in this study, as patients with high scores, suggesting advanced fibrosis, may warrant increased priority for HT. The limitations inherent to biopsy in a heterogenous process like CH suggest more comprehensive, multi-factorial evaluation of liver disease is sorely needed.

This study has several limitations. Given the specific population of interest, the study is limited by its small size and retrospective design introduces potential selection bias. Our analysis of post-HT survival is limited by the small number of patients with advanced fibrosis who underwent HT alone, as most patients found to have advanced fibrosis underwent CHLT. Further, while we relied on the most frequently utilized biopsy grading scale in CH, there is no universally accepting grading system. Additionally, the small number of patients necessitated grouping fibrosis stages (1 and 2, 3 and 4) for statistical analysis which may obscure between group differences for individual fibrosis stages. From a statistical standpoint, the small number of candidates with advanced fibrosis limited the number of variables that could be assessed in a multivariable model, requiring us to use a simulation-based approach. Finally, medications prior to transplant were not collected which may have influenced pre-HT laboratory studies.

In conclusion, here we show that a score based on three non-invasive factors reduced the need for LB during the assessment of HT candidates with congestive hepatopathy. We also found that LB fibrosis stage was not associated with post-HT survival and previously proposed clinical risk scores perform poorly in this population. The assessment of liver disease in patients with advanced HF remains a clinical challenge and requires further study in large, multi-center cohorts. Identification of novel strategies for the assessment of liver disease in this population stands to benefit the large group of patients whose transplant candidacy depends on the assessment of their liver disease.

## Data Availability

Please contact authors regarding data

## Acknowledgements

None

## Clinical Perspective

### What is new?

This study adds new data on the relationships between clinical variables (laboratory studies, transthoracic echocardiograms, and hemodynamics) and the development of liver fibrosis in patients with advanced heart failure. Further, we show that post-heart transplant survival in carefully selected patients does not differ by liver fibrosis grade. Finally, we propose a score of non-invasive factors which may help with evaluation of liver disease in heart transplant candidates.

### What are the clinical implications?

Evaluating liver disease in heart transplant candidates is difficult so there remains a reliance on liver biopsy. This study helps clinicians by elucidating relationships between other variables and liver fibrosis and shows that considering specific non-invasive factors (severe tricuspid regurgitation, alcohol use, and low-density lipoprotein) may reduce the need for liver biopsy.

Adding variables beyond the liver biopsy to aid in consideration of liver disease in heart transplant candidates is critical for the appropriate allocation of scarce organs.

## Non-standard Abbreviations and Acronyms

AAR-: AST to ALT ratio
AUROC-: area under the receiver operating characteristic curve
APRI –: AST to platelet ratio index
ALT-: alanine aminotransferase
AST-: aspartate aminotransferase
CH-: congestive hepatopathy
CHLT –: combined heart-liver transplant
EF-: ejection fraction
HT-: heart transplant LB – liver biopsy
LDL-: low-density lipoprotein
MELDXI-: Model for End Stage Liver Disease Excluding
INR MR-: mitral regurgitation
NRH-: nodular regenerative hyperplasia
OR-: odds ratio
RV-: right ventricular
TR-: tricuspid regurgitation
TTE-: transthoracic echocardiogram
VAST –: varices, ascites, splenomegaly, thrombocytopenia

---

**Supplementary Table 1.**
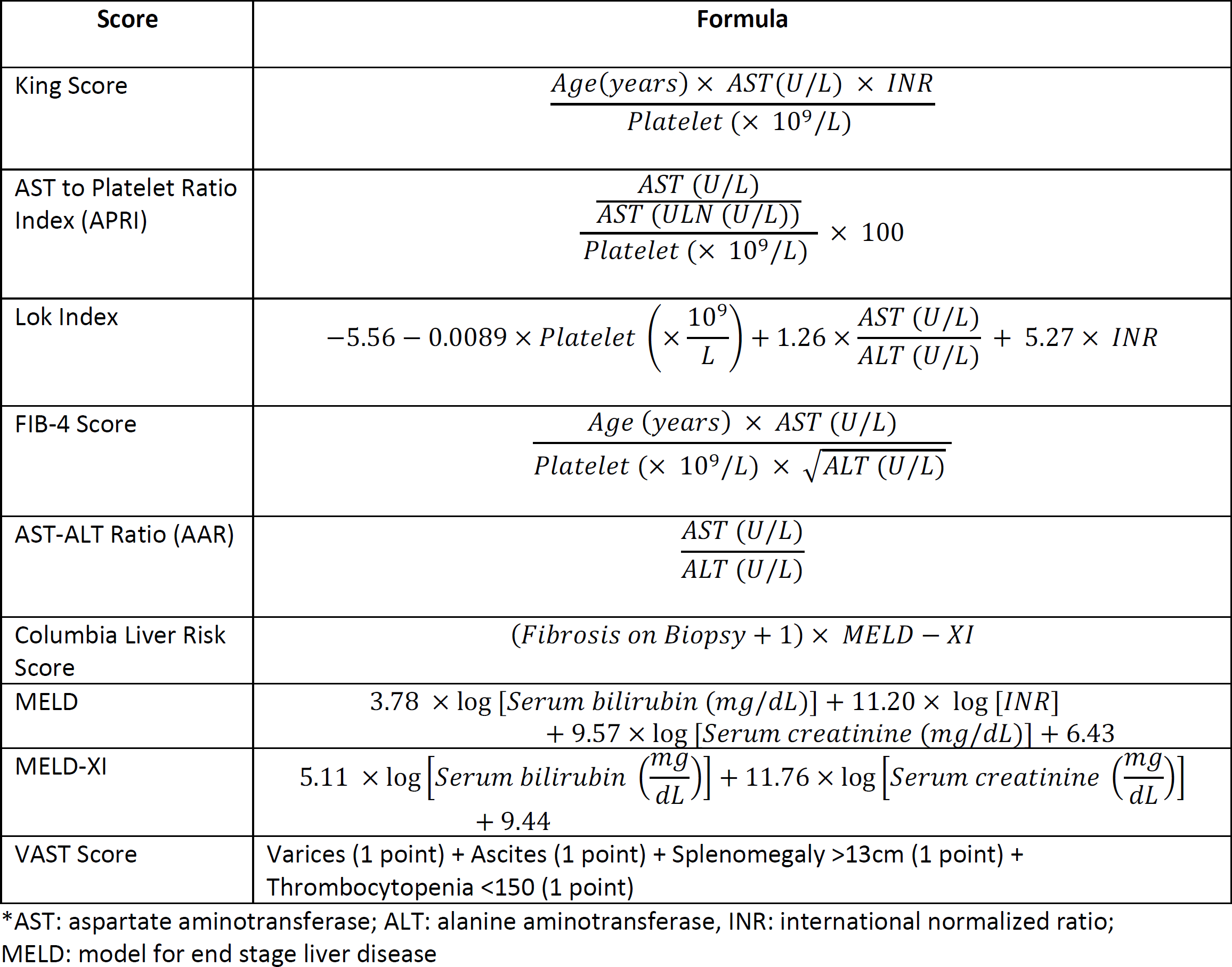
Risk Score Formulae.

**Supplementary Table 2.** Time (in days) between biological data and transplant or biopsy.

|  | Median Time (Days) | IQR |
| --- | --- | --- |
| Albumin (g/dL) | 16 | 8-46 |
| Total Bilirubin (mg/dL) | 89 | 48-238 |
| Serum Creatinine (mg/dL) | 38 | 18-130 |
| Platelet Count ($\times 10^9/L$) | 3 | 2-6 |
| Sodium (mmol/L) | 6 | 3-20 |
| Alanine Aminotransferase (units/L) | 152 | 43-912 |
| Aspartate Aminotransferase (units/L) | 55 | 26-443 |
| Transthoracic Echocardiogram | 42 | 16-156 |
| Right Heart Catheterization | 29 | 11-115 |

**Supplementary Table 3.** Clinical Risk Score for Advanced Fibrosis.

Cutoff for biopsy: Score $\geq -4$
| Variable | Points |
| --- | --- |
| Severe tricuspid regurgitation | 35 |
| Heavy alcohol use | 46 |
| LDL (minimum pre-transplant value ) | -1 * LDL value in mg/dL |

For example, for a candidate with severe tricuspid regurgitation (35 points), heavy alcohol use (46 point) and LDL of 65 (-65 points):

**Supplementary Table 4.** Missingness of Factors in Dataset.

| Factors | Missing |
| --- | --- |
| **Demographics** |  |
| Age | 0.00 |
| Gender | 0.00 |
| Etiology of Cardiomyopathy | 0.00 |
| Diabetes | 0.00 |
| Hypertension | 0.00 |
| BMI | 0.13 |
| Left Ventricular Assist Device | 0.00 |
| History of Heavy Alcohol Use | 0.00 |
| **Abdominal Ultrasound** |  |
| Ascites | 0.10 |
| Splenomegaly | 0.10 |
| Liver Parenchyma | 0.10 |
| Liver Size | 0.10 |
| **Transthoracic Echocardiogram** |  |
| Ejection Fraction | 0.06 |
| RV Function | 0.09 |
| Mitral Regurgitation | 0.08 |
| Tricuspid Regurgitation | 0.06 |
| LVIDD | 0.09 |
| TAPSE | 0.63 |
| IVC Compressibility | 0.14 |
| Right Atrial Pressure | 0.26 |
| Pulmonary Artery Systolic Pressure | 0.25 |
| Pulmonary Capillary Wedge Pressure | 0.27 |
| RAP to PCWP Ratio | 0.27 |
| **Laboratory Studies** |  |
| Albumin | 0.00 |
| Total Bilirubin | 0.00 |
| Serum Creatinine | 0.00 |
| Platelet Count | 0.00 |
| Sodium | 0.00 |
| Alanine Aminotransferase | 0.00 |
| Aspartate Aminotransferase | 0.00 |
| LDL | 0.00 |
| Triglycerides | 0.00 |
| Hepatitis B antibody | 0.00 |
| Hepatitis C antibody | 0.00 |
| **Non-Invasive Scores** |  |
| MELDXI | 0.00 |
| Lok Index | 0.00 |
| King Score | 0.00 |
| APRI | 0.00 |
| FIB4 | 0.00 |
| AAR | 0.00 |
| VAST | 0.00 |

Abbreviations: BMI- body mass index; LVIDD – left ventricular internal diameter end diastole; IVC – inferior vena cava; RAP- right atrium pressure; PCWP- pulmonary capillary wedge pressure

**Supplementary Table 5.** Univariable Analysis of Demographic, Imaging, Echocardiographic, Laboratory, and Hemodynamic Factors for Obtaining a Biopsy.

|  | Odds Ratio | 95% Confidence Intervals | p-value |
| --- | --- | --- | --- |
| **Demographics** |  |  |  |
| Age | 1.02 | 0.98-1.06 | 0.364 |
| Male | 1.12 | 0.40-3.10 | 0.829 |
| Non-Ischemic Cardiomyopathy | 2.27 | 0.88-5.86 | 0.089 |
| Diabetes | 0.54 | 0.21-1.40 | 0.207 |
| Hypertension | 0.82 | 0.35-1.91 | 0.643 |
| BMI | 0.99 | 0.91-1.08 | 0.850 |
| Left Ventricular Assist Device | 0.98 | 0.92-1.03 | 0.371 |
| History of heavy alcohol use | 1.91 | 0.86-4.22 | 0.112 |
| Hepatitis C Positive | 1.40 | 0.37-5.34 | 0.620 |
| Hepatitis B Positive | 0.48 | 0.20-1.15 | 0.100 |
| **Abdominal Ultrasound** |  |  |  |
| Ascites | 4.19 | 1.69-10.42 | 0.002 |
| Splenomegaly | 2.16 | 0.90-5.18 | 0.084 |
| Liver Parenchyma |  |  |  |
| Heterogenous | 3.64 | 1.06-12.54 | 0.041 |
| Nodular | 7.48 | 2.33-24.01 | 0.001 |
| Liver Size |  |  |  |
| Large | 1.95 | 0.83-4.56 | 0.123 |
| **Transthoracic Echocardiogram** |  |  |  |
| Ejection Fraction | 1.01 | 0.98-1.04 | 0.538 |
| RV Function |  |  |  |
| Moderately depressed | 0.72 | 0.21-2.46 | 0.599 |
| Severely depressed | 1.41 | 0.43-4.63 | 0.573 |
| Mitral Regurgitation |  |  |  |
| Moderate | 2.87 | 0.75-11.06 | 0.125 |
| Severe | 3.91 | 1.03-14.88 | 0.046 |
| Tricuspid Regurgitation |  |  |  |
| Moderate | 0.83 | 0.16-4.27 | 0.827 |
| Severe | 5.88 | 1.63-21.19 | 0.007 |
| TAPSE | 1.74 | 0.35-8.56 | 0.497 |
| IVC Compressibility |  |  |  |
| None | 1.87 | 0.69-5.06 | 0.218 |
| **Hemodynamics** |  |  |  |
| Right Atrial Pressure | 1.09 | 1.02-1.15 | 0.010 |
| Pulmonary Artery Systolic Pressure | 0.98 | 0.94-1.01 | 0.180 |
| Pulmonary Capillary Wedge Pressure | 1.00 | 0.95-1.05 | 0.882 |
| RAP to PCWP Ratio | 6.44 | 1.56-26.66 | 0.010 |
| **Laboratory Studies** |  |  |  |
| Albumin | 1.77 | 0.83-3.76 | 0.139 |
| Total Bilirubin | 0.53 | 0.14-1.98 | 0.348 |
| Serum Creatinine | 1.59 | 0.94-2.68 | 0.081 |
| Platelet Count | 1.00 | 0.99-1.01 | 0.751 |
| Sodium | 0.97 | 0.89-1.06 | 0.483 |
| Alanine Aminotransferase | 0.98 | 0.93-1.03 | 0.378 |
| Aspartate Aminotransferase | 0.98 | 0.93-1.02 | 0.311 |
| LDL | 1.00 | 0.99-1.99 | 0.501 |
| Triglycerides | 1.00 | 0.99-1.00 | 0.090 |
| **Non-Invasive Scores** |  |  |  |
| MELDIXI | 1.05 | 0.94-1.17 | 0.426 |
| Lok Index | 1.04 | 0.83-1.31 | 0.724 |
| King Score | 0.98 | 0.95-1.01 | 0.224 |
| APRI | 0.51 | 0.21-1.24 | 0.137 |
| FIB4 | 0.74 | 0.36-1.49 | 0.396 |
| AAR | 1.88 | 0.76-4.68 | 0.174 |
| VAST | 2.87 | 1.97-4.18 | <0.001 |

Abbreviations: BMI- body mass index; IVC – inferior vena cava; RAP- right atrium pressure ; PCWP- pulmonary capillary wedge pressure; APRI- AST to platelet ratio index; AAR- AST to ALT ratio; MELDXI- model for end stand liver disease excluding INR; VAST- varices, ascites, splenomegaly, thrombocytopenia

**Supplementary Figure 1.**
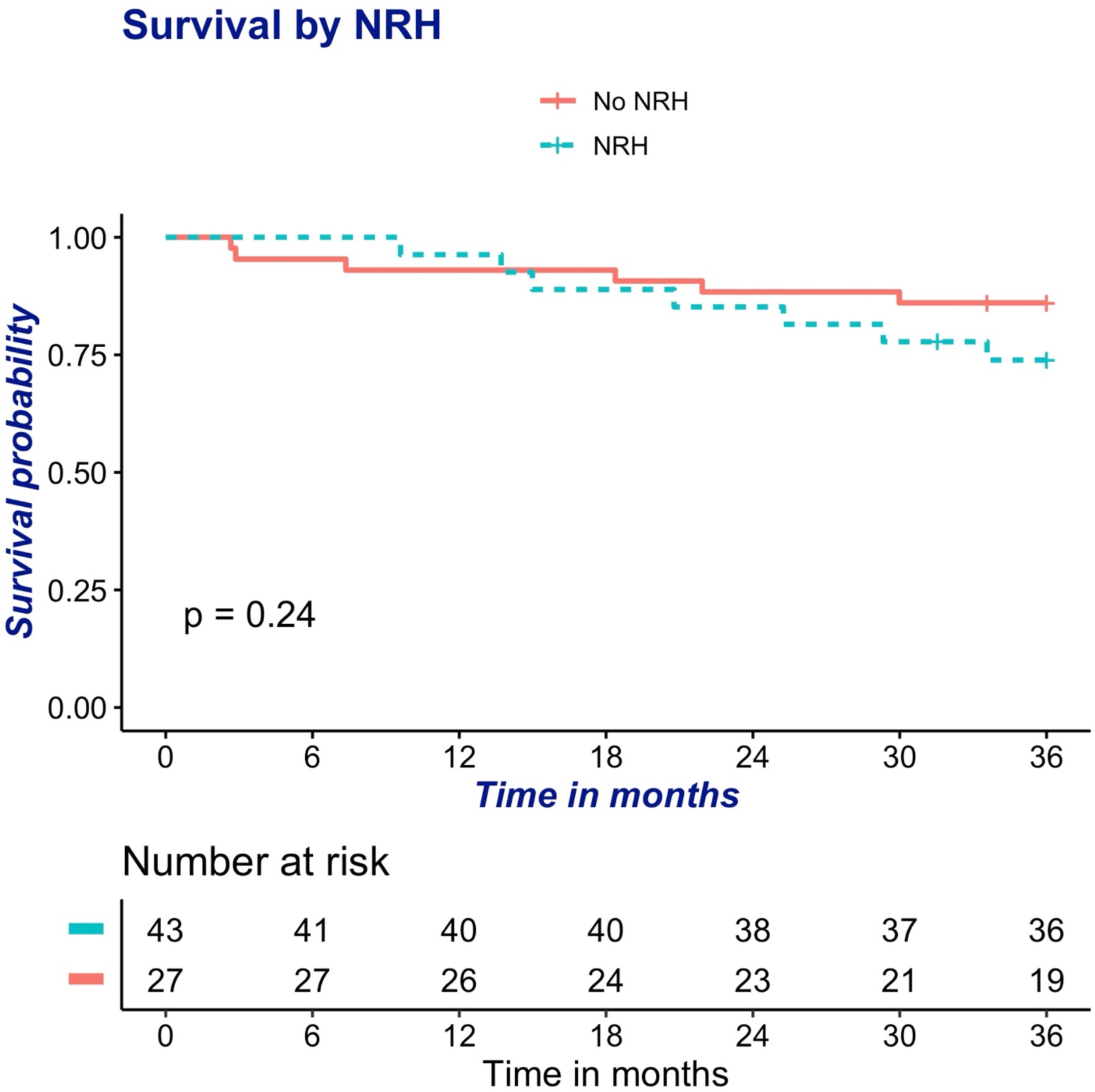
Survival based on NRH presence on liver biopsy.

**Supplementary Figure 2.**
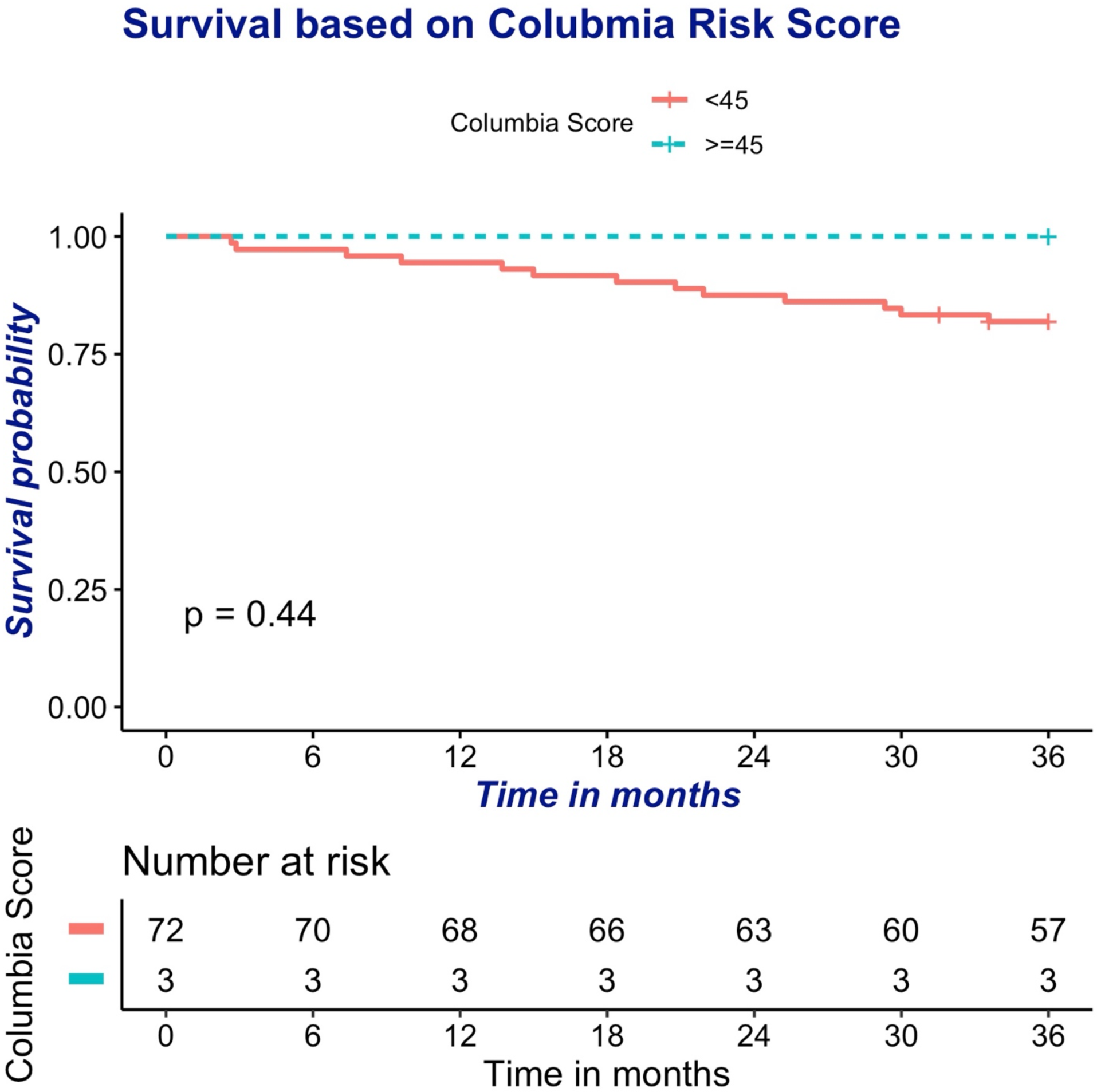
Survival based on Columbia Risk Score.

**Supplementary Table 6.**
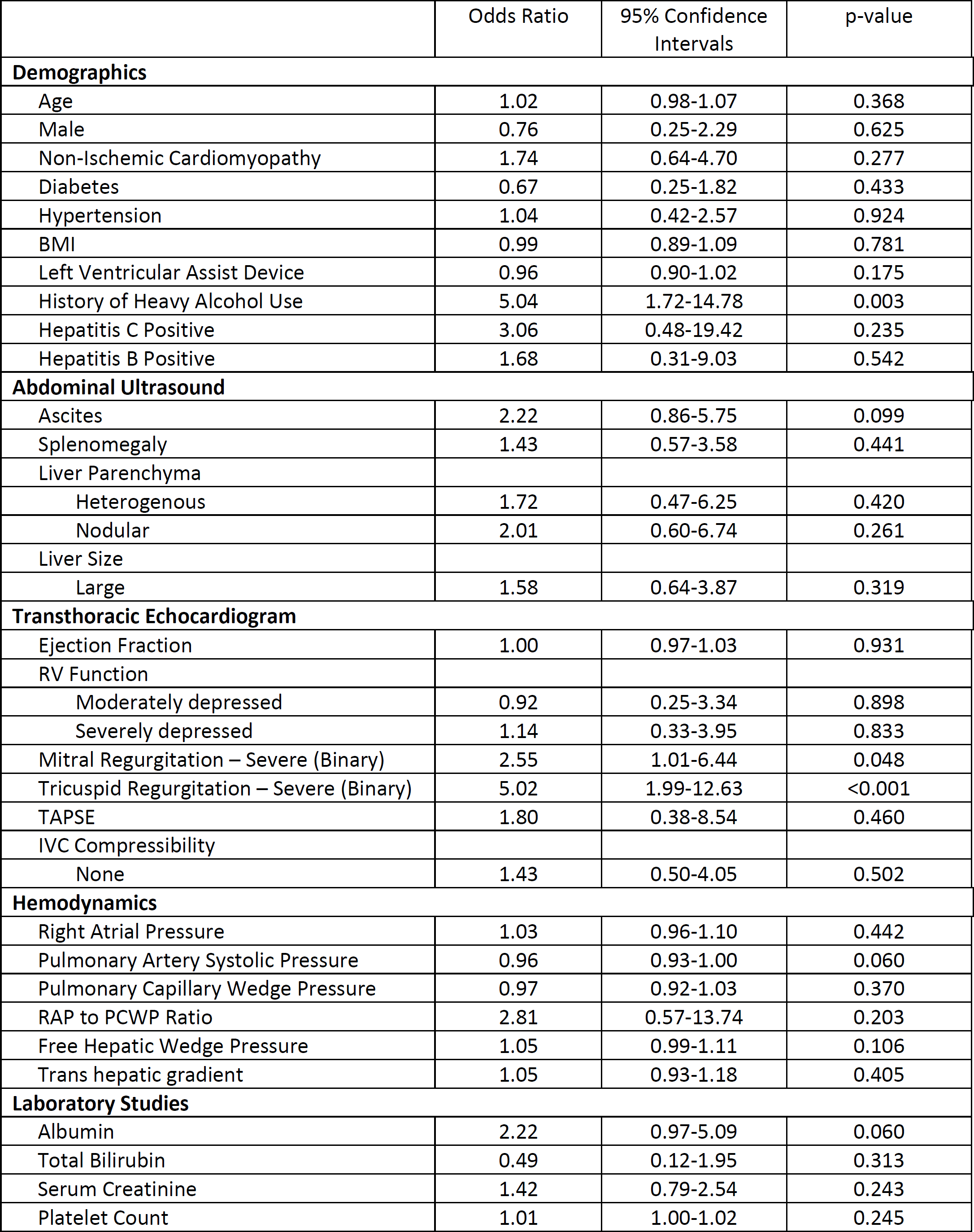
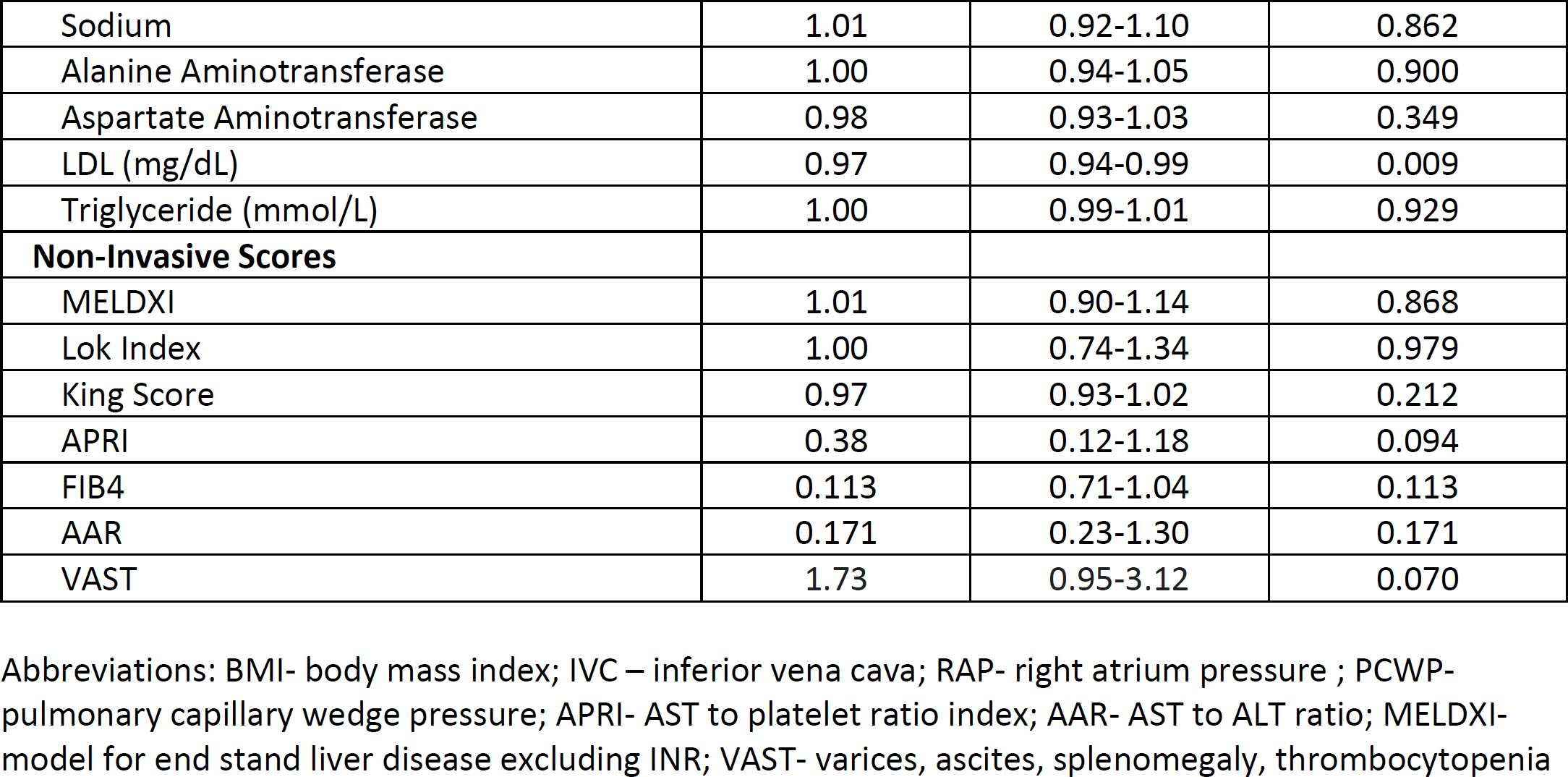
Univariable Analysis of Demographic, Imaging, Echocardiographic, Laboratory, and Hemodynamic Factors for Advanced Fibrosis.

|  | Odds Ratio | 95% Confidence Intervals | p-value |
| --- | --- | --- | --- |
| **Demographics** |  |  |  |
| Age | 1.02 | 0.98-1.07 | 0.368 |
| Male | 0.76 | 0.25-2.29 | 0.625 |
| Non-Ischemic Cardiomyopathy | 1.74 | 0.64-4.70 | 0.277 |
| Diabetes | 0.67 | 0.25-1.82 | 0.433 |
| Hypertension | 1.04 | 0.42-2.57 | 0.924 |
| BMI | 0.99 | 0.89-1.09 | 0.781 |
| Left Ventricular Assist Device | 0.96 | 0.90-1.02 | 0.175 |
| History of Heavy Alcohol Use | 5.04 | 1.72-14.78 | 0.003 |
| Hepatitis C Positive | 3.06 | 0.48-19.42 | 0.235 |
| Hepatitis B Positive | 1.68 | 0.31-9.03 | 0.542 |
| **Abdominal Ultrasound** |  |  |  |
| Ascites | 2.22 | 0.86-5.75 | 0.099 |
| Splenomegaly | 1.43 | 0.57-3.58 | 0.441 |
| Liver Parenchyma |  |  |  |
| Heterogenous | 1.72 | 0.47-6.25 | 0.420 |
| Nodular | 2.01 | 0.60-6.74 | 0.261 |
| Liver Size |  |  |  |
| Large | 1.58 | 0.64-3.87 | 0.319 |
| **Transthoracic Echocardiogram** |  |  |  |
| Ejection Fraction | 1.00 | 0.97-1.03 | 0.931 |
| RV Function |  |  |  |
| Moderately depressed | 0.92 | 0.25-3.34 | 0.898 |
| Severely depressed | 1.14 | 0.33-3.95 | 0.833 |
| Mitral Regurgitation – Severe (Binary) | 2.55 | 1.01-6.44 | 0.048 |
| Tricuspid Regurgitation – Severe (Binary) | 5.02 | 1.99-12.63 | <0.001 |
| TAPSE | 1.80 | 0.38-8.54 | 0.460 |
| IVC Compressibility |  |  |  |
| None | 1.43 | 0.50-4.05 | 0.502 |
| **Hemodynamics** |  |  |  |
| Right Atrial Pressure | 1.03 | 0.96-1.10 | 0.442 |
| Pulmonary Artery Systolic Pressure | 0.96 | 0.93-1.00 | 0.060 |
| Pulmonary Capillary Wedge Pressure | 0.97 | 0.92-1.03 | 0.370 |
| RAP to PCWP Ratio | 2.81 | 0.57-13.74 | 0.203 |
| Free Hepatic Wedge Pressure | 1.05 | 0.99-1.11 | 0.106 |
| Trans hepatic gradient | 1.05 | 0.93-1.18 | 0.405 |
| **Laboratory Studies** |  |  |  |
| Albumin | 2.22 | 0.97-5.09 | 0.060 |
| Total Bilirubin | 0.49 | 0.12-1.95 | 0.313 |
| Serum Creatinine | 1.42 | 0.79-2.54 | 0.243 |
| Platelet Count | 1.01 | 1.00-1.02 | 0.245 |
| Sodium | 1.01 | 0.92-1.10 | 0.862 |
| Alanine Aminotransferase | 1.00 | 0.94-1.05 | 0.900 |
| Aspartate Aminotransferase | 0.98 | 0.93-1.03 | 0.349 |
| LDL (mg/dL) | 0.97 | 0.94-0.99 | 0.009 |
| Triglyceride (mmol/L) | 1.00 | 0.99-1.01 | 0.929 |
| **Non-Invasive Scores** |  |  |  |
| MELDIXI | 1.01 | 0.90-1.14 | 0.868 |
| Lok Index | 1.00 | 0.74-1.34 | 0.979 |
| King Score | 0.97 | 0.93-1.02 | 0.212 |
| APRI | 0.38 | 0.12-1.18 | 0.094 |
| FIB4 | 0.113 | 0.71-1.04 | 0.113 |
| AAR | 0.171 | 0.23-1.30 | 0.171 |
| VAST | 1.73 | 0.95-3.12 | 0.070 |

Abbreviations: BMI- body mass index; IVC – inferior vena cava; RAP- right atrium pressure ; PCWP- pulmonary capillary wedge pressure; APRI- AST to platelet ratio index; AAR- AST to ALT ratio; MELDXI- model for end stand liver disease excluding INR; VAST- varices, ascites, splenomegaly, thrombocytopenia

